# Respiratory virus co-infection is a risk factor for worse outcomes during *Staphylococcus aureus* bacteraemia

**DOI:** 10.1101/2025.10.28.25338947

**Authors:** Katherine Roberts, Simon Dewar, Rebecca K. Sutherland, Clark D. Russell

## Abstract

We conducted a retrospective study of adults with *Staphylococcus aureus* bacteraemia to determine the impact of respiratory virus co-infection. Co-infection, predominantly with SARS-CoV-2, was associated with SAB originating from the respiratory tract (21.1% vs. 5.2%, *P*=0.002), persistent bacteraemia (13.2% *vs*. 3.4%, *P*=0.02) and independently with 30-day mortality (31.6% *vs*. 18.0%, *P*=0.04).

## INTRODUCTION

Respiratory virus infection in humans can be complicated by secondary bacterial pneumonia caused by gram-positive cocci, including *Staphylococcus aureus*, associated with increased mortality[1-5]. Mouse models of respiratory virus/*S. aureus* co-infection demonstrate that viral infection lowers the threshold for asymptomatic *S. aureus* nasal colonisation to progress to pneumonia[4, 6]. Mechanistically, influenza A virus infection impairs phagocyte NADPH oxidase activity and pulmonary interleukin 17 production in mice in response to *S. aureus*[7, 8]. These factors are both key elements of the host response to *S. aureus*[9]. We therefore reasoned that respiratory virus infection could impact on the outcome of *S. aureus* bacteraemia (SAB) and aimed to test the hypothesis that respiratory virus co-infection was associated with worse clinical outcomes in SAB.

## METHODS

We analysed data from an ongoing retrospective cohort study of consecutive adults (aged ≥18 years) with monomicrobial SAB in the United Kingdom (Edinburgh and the Lothians), as previously described[10]. This study is approved by the South East Scotland Research Ethics Committee 02 (23/SS/0025/AM02). Patients were identified through a laboratory database search for all blood cultures with growth of *S. aureus* from 8^th^ January 2021 to 29^th^ December 2024. These dates were chosen to avoid inclusion of large numbers of people with Covid-19 prior to the availability of vaccination and therapeutics. Results of respiratory virus testing performed between 7 days before to 3 days after the index positive blood culture were included. Respiratory virus testing was done using either point of care or routine laboratory-based testing. Point of care analysers used were Cepheid GeneXpert, Roche Liat, and Roche Eplex. Laboratory analysers used were Alinity M (Abbott) and Seegene (Mast). All-cause mortality was recorded 30 days after the index positive blood culture. Persistent SAB was defined as a positive blood culture for *S. aureus* >48h after the index blood culture whilst still receiving treatment.

Variables were compared between patients tested/not tested for respiratory viruses, then between patients with/without confirmed co-infection. Categorical variables were compared using Fisher’s Exact or Chi-squared tests and continuous variables were compared using Mann Whitney U or Kruskal-Wallis tests with correction for multiple testing where appropriate. Survival was compared using Kaplan Meir curves and the log rank test. Multiple logistic regression was used to identify independent risk factors for mortality. R (Version 4.2.2) and GraphPad Prism Version 10.5.0 for macOS were used for data analysis and visualisation.

## RESULTS

We identified 651 patients with SAB during the study period (**Supplementary Figure 1**). 64.5% (n=420/651) underwent testing for SARS-CoV-2, influenza A, influenza B, and respiratory syncytial virus (RSV). 38 of these 420 tested patients (9.1%) tested positive for a respiratory virus. 30 tested positive for SARS-CoV-2, seven tested positive for influenza A, and one tested positive for RSV.

### Clinical characteristics

There were no differences in most baseline patient characteristics between those tested for respiratory viruses and those not tested, including age, sex, Charlson Comorbidity Index (CCI), and qSOFA score (**Supplementary Table 1**). Consistent with enhanced infection prevention and control practices in haemodialysis units, patients undergoing haemodialysis were more likely to be tested. People who inject drugs were less likely to be tested. The portal of entry of bacteraemia also differed between tested/not tested patients, with an intravenous catheter and the respiratory tract both more common amongst patients undergoing testing.

Baseline patient characteristics also did not differ significantly between patients with/without a co-infection detected, including age, sex, CCI, and qSOFA score (**Supplementary Table 2**). Patients with co-infection were substantially more likely to have SAB originating from the respiratory tract (21.1% vs. 5.2%, *P*=0.002). There was a non-significant trend towards an association between co-infection and healthcare related acquisition (**Supplementary Table 2**).

### Outcomes

There was no significant difference in the incidence of metastatic complications between patients with/without co-infection but patients with co-infection were more likely to have persistent bacteraemia (13.2% vs. 3.4%, *P* = 0.02). 30-day all-cause mortality was also higher amongst patients with co-infection (31.6% vs. 18.0%, *P* = 0.04) with the difference mainly driven by deaths within the first 10 days since the index blood culture (**Figure 1A**). Respiratory virus co-infection remained independently associated with mortality after adjusting for age, sex, and co-morbidity (odds ratio = 2.3, 95% confidence interval 1.03-5.05, *P* = 0.04; **Figure 1B**). 30-day mortality and persistent bacteraemia represented largely distinct outcomes and this could reflect the competing risk of death when assessing persistent bacteraemia (**Figure 1C**).

**Figure 1.**
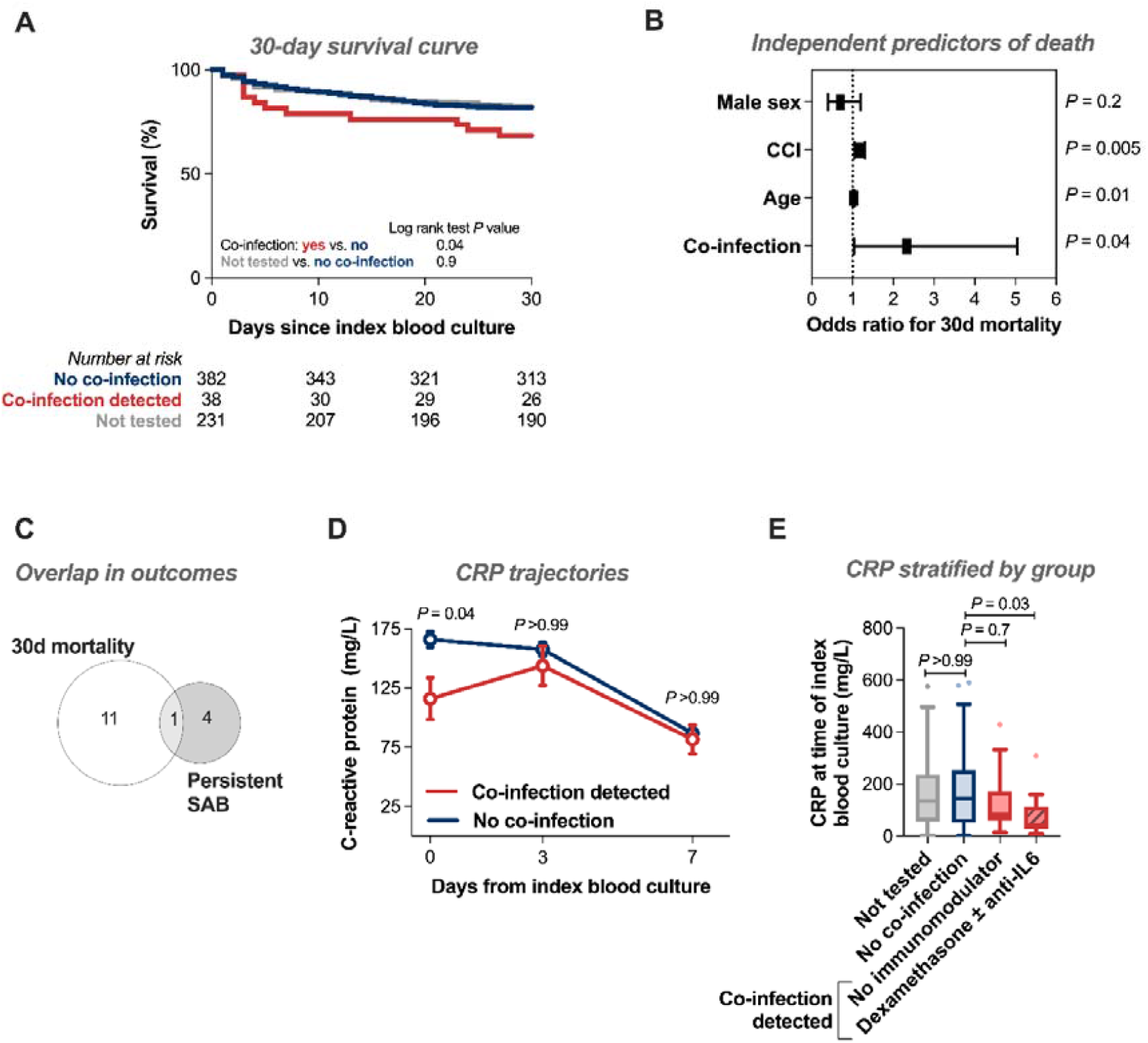
Respiratory virus co-infection during *Staphylococcus aureus* bacteraemia. (A)Kaplan-Meier survival curve for unadjusted 30-day survival. Groups compared using the log-rank test. **(B)** Results of multiple logistic regression with 30-day mortality as outcome. Graph shows odds ratio and 95% confidence interval. **(C)** Euler diagram representing overlap in number of patients who died within 30 days and who were diagnosed with persistent SAB. **(D)** Line graph representing CRP concentration on days 0, 3, and 7 from index blood culture. Graph shows mean and standard error of the mean for each timepoint. Groups compared using Kruskal-Wallis test with Dunn’s test for multiple comparisons. **(E)** Box and whisker plot drawn using Tukey’s method. Box shows interquartile range and horizontal line shows median. Groups compared using Kruskal-Wallis test with Dunn’s test for multiple comparisons.

### Impact of Covid-19 immunomodulatory therapies

C-reactive protein (CRP) concentration at the time of index blood culture (day zero) was lower in patients with co-infection then increased to a similar concentration by day three, appearing to represent a lag (**Figure 1D**). There was no difference in the day zero total white blood cell count (**Supplementary Figure 2**). Of 30 patients with SARS-CoV-2 co-infection, 11 received dexamethasone alone and 1 received dexamethasone plus tocilizumab. The lowest median day zero CRP was seen in patients receiving these Covid-19 immunomodulatory treatments though there was still a trend of lower CRP in the patients with co-infection not receiving these treatments (**Figure 1E**). Receipt of immunomodulatory therapy did not increase the likelihood of 30-day mortality or persistent SAB (**Supplementary Table 3**).

## DISCUSSION

Respiratory virus co-infection during SAB was associated with an increased risk of early death and persistent bacteraemia. These differences were not explained by differences in baseline characteristics, such as age, sex, or co-morbidity, nor receipt of Covid-19 immunomodulatory therapies.

Our study has some important limitations. Our findings are from a single centre requiring validation in additional cohorts to ensure generalisability. Only 64.5% of patients underwent testing for respiratory virus co-infection, likely based on clinical suspicion, meaning that there will be a selection bias. The sample size of patients who tested positive was small (n= 38), reducing statistical power, and the findings predominantly relate to SARS-CoV-2 co-infection. There is a competing risk between mortality and persistent bacteraemia, meaning that patients who died early may not have had the opportunity to be diagnosed with persistent SAB, potentially underestimating the association between coinfection and persistent bacteraemia. Time to receipt of antimicrobials active against *S. aureus* was not recorded, and it is possible that delayed receipt of antimicrobials in patients with confirmed/suspected respiratory virus infection could also explain the worse outcomes in this group.

Based on observations from mouse models[4, 6-8] we hypothesise that the worse outcomes in the co-infected group relate to virus-mediated interference with host defence, resulting in impaired antibacterial responses which could represent treatable traits. Impaired *ex vivo* neutrophil phagocytosis has been identified in adults with critical illness of any cause, which can be reversed with GM-CSF[11]. Similarly, interferon gamma has been shown to improve monocyte dysfunction in adults with septic shock[12]. Future work should investigate host responses during *S. aureus*/respiratory virus co-infection in humans to determine if the observed deficits (e.g. NADPH oxidase, IL-17) described in mice also apply to humans and could therefore represent targets for host-directed therapies. In conclusion, clinical management of patients with SAB and respiratory virus co-infection requires optimisation to improve on current outcomes. Such strategies could have broader utility in the setting of secondary bacterial pneumonia following respiratory virus infection.

## Data Availability

All data produced in the present work are contained in the manuscript

## Funding sources

CDR was supported by the Chief Scientist Office (CSO) Scotland (PCL/25/21). For the purpose of open access, the author has applied a creative commons attribution (CC BY) licence to any author accepted manuscript version arising.

## Potential conflicts of interest

None

## Author contributions

CDR conceived the study. KR and CDR curated the data, did the investigation, did the formal analysis, and had access to the raw data. CDR supervised the study and validated the study data. KR wrote the original draft of the manuscript. All authors reviewed and edited the manuscript. The corresponding author had full access to all the data and final responsibility for the decision to submit for publication.

## Data sharing

Please contact the corresponding author to discuss. We would welcome opportunities to share data and contribute to collaborative analyses.

## Acknowledgements

This work used data provided by patients and collected by the NHS as part of their care.

## Notes

### Competing Interest Statement

The authors have declared no competing interest.

### Author Declarations

This study is approved by the South East Scotland Research Ethics Committee 02 (23/SS/0025/AM02).

## REFERENCES

1. Morens DM, Taubenberger JK, Fauci AS. Predominant role of bacterial pneumonia as a cause of death in pandemic influenza: implications for pandemic influenza preparedness. J Infect Dis 2008; 198(7): 962–70.

2. Morris DE, Cleary DW, Clarke SC. Secondary Bacterial Infections Associated with Influenza Pandemics. Front Microbiol 2017; 8: 1041.

3. Russell CD, Fairfield CJ, Drake TM, et al. Co-infections, secondary infections, and antimicrobial use in patients hospitalised with COVID-19 during the first pandemic wave from the ISARIC WHO CCP-UK study: a multicentre, prospective cohort study. Lancet Microbe 2021; 2(8): e354–e65.

4. Lubkin A, Bernard-Raichon L, DuMont AL, et al. SARS-CoV-2 infection predisposes patients to coinfection with Staphylococcus aureus. mBio 2024; 15(8): e0166724.

5. Gerver SM, Guy R, Wilson K, et al. National surveillance of bacterial and fungal coinfection and secondary infection in COVID-19 patients in England: lessons from the first wave. Clin Microbiol Infect 2021; 27(11): 1658–65.

6. Reddinger RM, Luke-Marshall NR, Hakansson AP, Campagnari AA. Host Physiologic Changes Induced by Influenza A Virus Lead to Staphylococcus aureus Biofilm Dispersion and Transition from Asymptomatic Colonization to Invasive Disease. mBio 2016; 7(4).

7. Sun K, Metzger DW. Influenza infection suppresses NADPH oxidase-dependent phagocytic bacterial clearance and enhances susceptibility to secondary methicillin-resistant Staphylococcus aureus infection. J Immunol 2014; 192(7): 3301–7.

8. Kudva A, Scheller EV, Robinson KM, et al. Influenza A inhibits Th17-mediated host defense against bacterial pneumonia in mice. J Immunol 2011; 186(3): 1666–74.

9. Russell CD, Goeldner-Thompson S, Smith E, et al. Genome-Scale Meta-analysis of Host Responses to Staphylococcus aureus Identifies Pathways for Host-Directed Therapeutic Targeting. J Infect Dis 2025; 232(2): e290–e300.

10. Russell CD, Berry K, Cooper G, et al. Distinct Clinical Endpoints of Staphylococcus aureus Bacteraemia Complicate Assessment of Outcome. Clin Infect Dis 2024; 79(3): 604–11.

11. Pinder EM, Rostron AJ, Hellyer TP, et al. Randomised controlled trial of GM-CSF in critically ill patients with impaired neutrophil phagocytosis. Thorax 2018; 73(10): 918–25.

12. Döcke W-D, Randow F, Syrbe U, et al. Monocyte deactivation in septic patients: Restoration by IFN-γ treatment. Nature Medicine 1997; 3(6): 678–81.

