## Supplementary materials for "Respiratory virus co-infection is a risk factor for worse outcomes during *Staphylococcus aureus* bacteraemia"

Supplementary material

### Supplementary Table 1: Baseline characteristics for participants tested vs. not tested for respiratory viruses

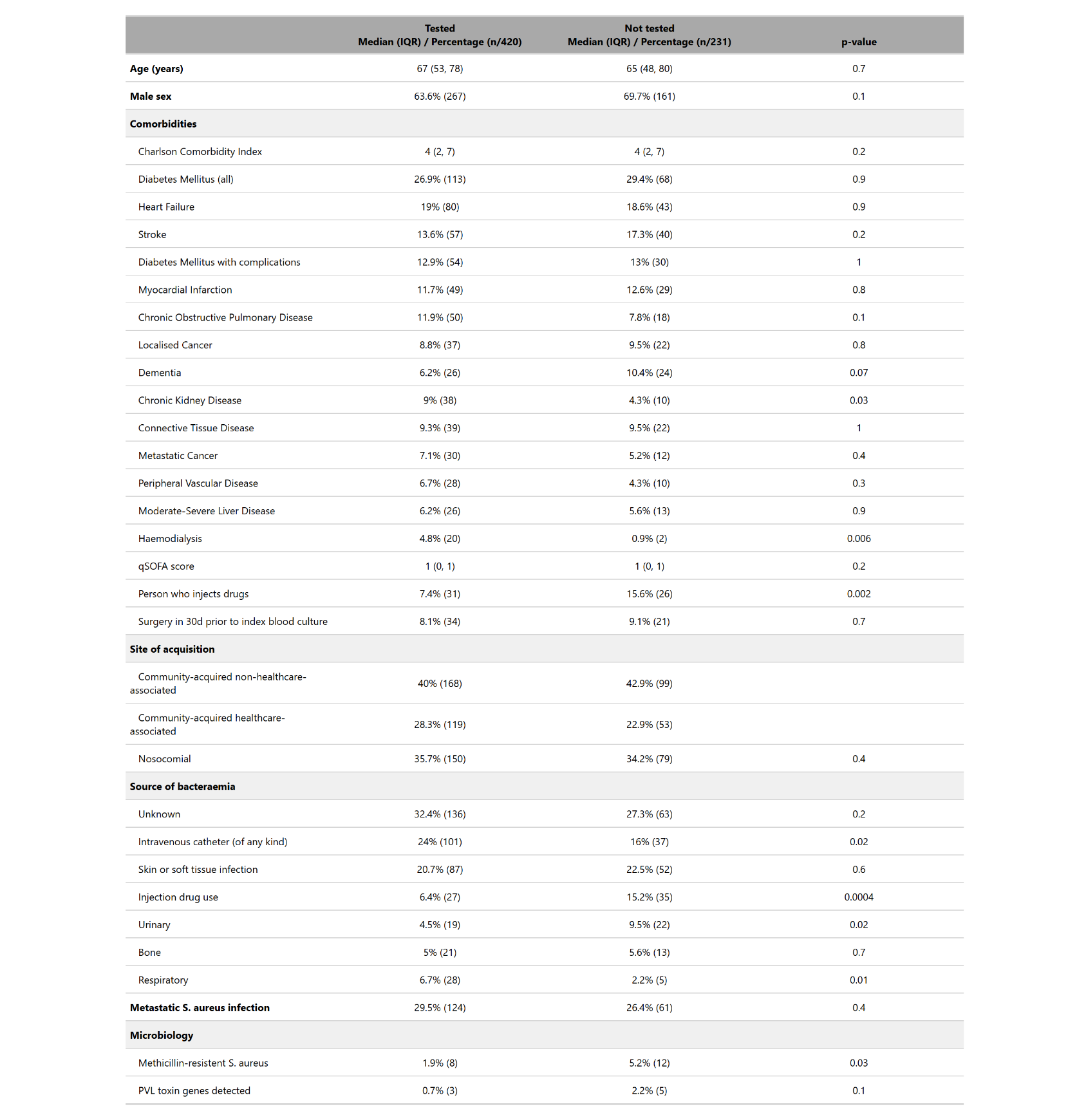
Categorical variables were compared using Fisher’s Exact or Chi-squared tests and continuous variables were compared using the Mann Whitney U test. Missing data were removed for qSOFA score (47 missing for participants not tested and 71 missing for tested) and for PVL toxin genes detected (4 missing for not tested participants and 8 missing for tested).

### Supplementary Table 2: Baseline characteristics for participants tested positive vs. tested negative for respiratory viruses

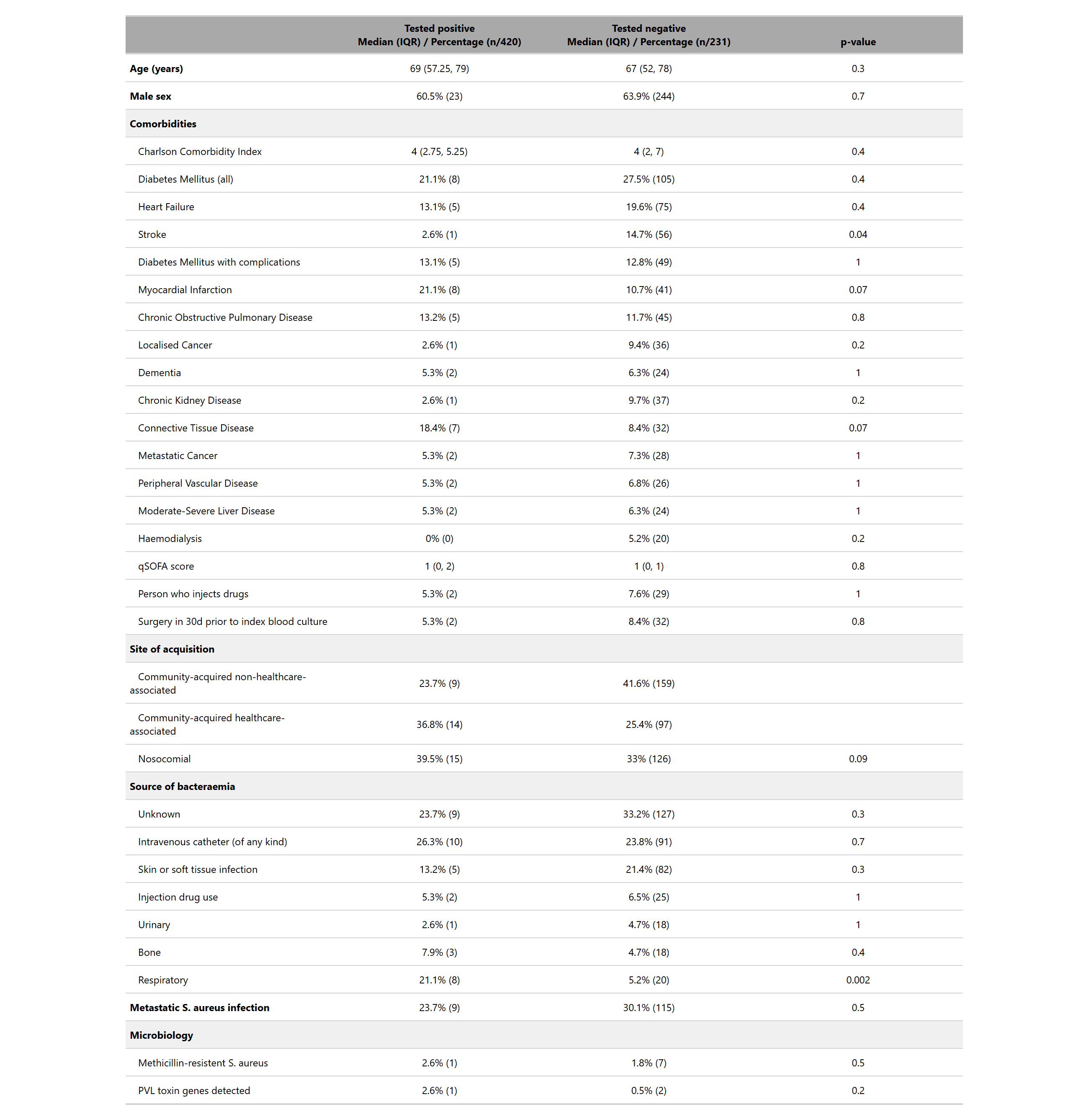

Categorical variables were compared using Fisher’s Exact or Chi-squared tests and continuous variables were compared using the Mann Whitney U test. Missing data were removed for qSOFA score (5 missing for positive participants and 47 missing for negative participants) and for PVL toxin genes detected (2 missing for positive participants and 6 missing for negative participants).

### Supplementary Table 3: Impact of Covid-19 immunomodulatory therapies

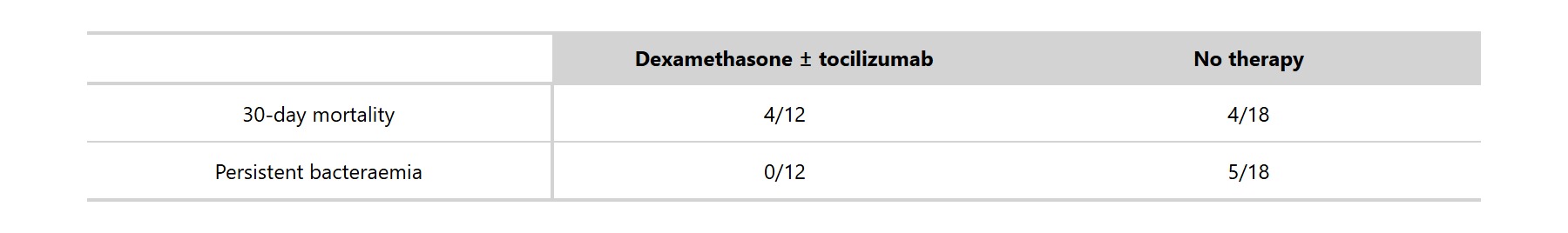

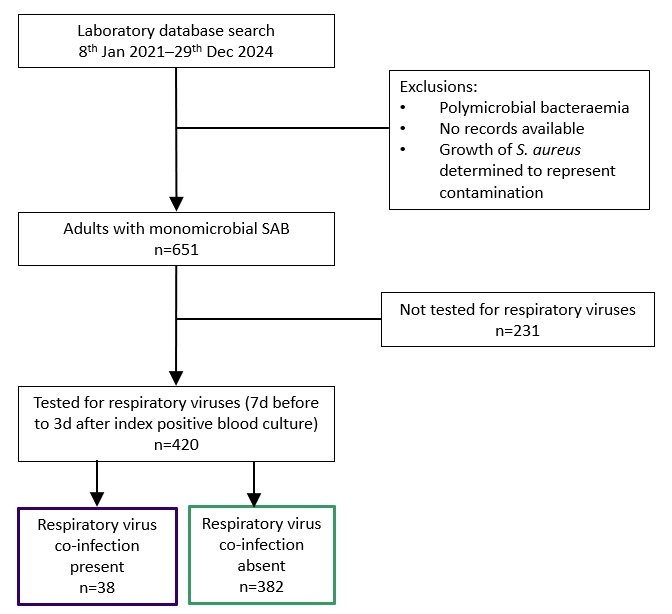

### Supplementary Figure 1: Flow diagram for Edinburgh retrospective observational cohort

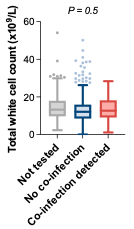

### Supplementary Figure 2: Total white cell count at time of index blood culture

Box and whisker plot drawn using Tukey’s method. Box shows interquartile range and horizontal line shows median. Groups compared using Mann Whitney test.
